# Tissue contamination challenges the credibility of machine learning models in real world digital pathology

**DOI:** 10.1101/2023.04.28.23289287

**Authors:** Ismail Irmakci, Ramin Nateghi, Rujoi Zhou, Ashley E. Ross, Ximing J. Yang, Lee A. D. Cooper, Jeffery A. Goldstein

## Abstract

Machine learning (ML) models are poised to transform surgical pathology practice. The most successful use attention mechanisms to examine whole slides, identify which areas of tissue are diagnostic, and use them to guide diagnosis. Tissue contaminants, such as floaters, represent unexpected tissue. While human pathologists are extensively trained to consider and detect tissue contaminants, we examined their impact on ML models.

We trained 4 whole slide models. Three operate in placenta for 1) detection of decidual arteriopathy (DA), 2) estimation of gestational age (GA), and 3) classification of macroscopic placental lesions. We also developed a model to detect prostate cancer in needle biopsies. We designed experiments wherein patches of contaminant tissue are randomly sampled from known slides and digitally added to patient slides and measured model performance. We measured the proportion of attention given to contaminants and examined the impact of contaminants in T-distributed Stochastic Neighbor Embedding (tSNE) feature space.

Every model showed performance degradation in response to one or more tissue contaminants. DA detection balanced accuracy decreased from 0.74 to 0.69 +/- 0.01 with addition of 1 patch of prostate tissue for every 100 patches of placenta (1% contaminant). Bladder, added at 10% contaminant raised the mean absolute error in estimating gestation age from 1.626 weeks to 2.371 +/ 0.003 weeks. Blood, incorporated into placental sections, induced false negative diagnoses of intervillous thrombi. Addition of bladder to prostate cancer needle biopsies induced false positives, a selection of high-attention patches, representing 0.033mm^2^, resulted in a 97% false positive rate when added to needle biopsies. Contaminant patches received attention at or above the rate of the average patch of patient tissue.

Tissue contaminants induce errors in modern ML models. The high level of attention given to contaminants indicates a failure to encode biological phenomena. Practitioners should move to quantify and ameliorate this problem.

## INTRODUCTION

Machine learning (ML) models are poised to transform pathology practice. Models have been developed to detect and grade cancers, quantify immunohistochemistry, and identify transplant rejection (1–5). The most successful and potentially transformative are those that take entire slides as input and produce slide or patient-level diagnoses (6–10). To process the large amount of data represented by each slide, these models can rely on attention or other pooling mechanisms to identify key areas on the slide. The attention is then used to explain the model’s decision and guide pathologists to areas of concerns. Any ML model can give unpredictable results when presented with out-of-distribution data not seen during training, but the impact on attention is less studied. Pathology also presents a unique challenge to attention-based models because of the sporadic presence of tissue contaminants – material from different patients or specimens that is unintentionally included in the slide (11). The goal of this study is to test the impact of tissue contaminants on model performance and examine how tissue contaminants interact with the attention mechanism.

### Tissue contamination

The process of tissue handling, wherein patient tissue becomes a slide, contains multiple steps in which tissues from one patient can appear on the slide of a different patient. This could be a “push” from an insufficiently cleaned tool at the grossing bench, “block contamination” that occurs during transport processing in a retort shared by tissues from multiple patients, or a “floater” that occurs when histology water baths are insufficiently cleaned between blocks (11–14). These errors, collectively referred to as “tissue contamination” are well described in the pathology literature, but often come as a surprise to non-pathologist researchers or physicians (15).

Tissue contaminants have been identified in up to 3% of slides examined with an average size of 1 mm^2^ (11,13). 12.7% of contaminants appeared to be neoplasia and 0.4% were judged to have a high risk of causing misdiagnosis. This low proportion belies a surprising count. An institution that produces 1 million slides per year (ours does), can expect 30,000 slides with tissue contaminants, of which 120 have a high risk of causing error. Nonetheless, the actual error rate may be much lower. In a review of 276 legal cases against pathologists for misdiagnosis 2004-2010, only one involved a floater (16).

### Human learning, machine learning

The low error rate due to tissue contaminants in real world clinical practice may relate to pathologist education. Demonstrated knowledge of normal histology is a basic level 1 (of 5) milestone competency in pathology resident education in the U.S. (17,18). Identification of specimen integrity issues, specifically including floaters, is a level 2 skill (17,18). Thus, any pathologist examining a placental slide with tissue contamination by prostate should 1) identify which portions of the specimen are placenta and which are prostate and 2) recognize that the prostate tissue is present in error and ignore it.

The contrast with machine learning approaches is stark. Machine learning approaches in digital pathology generally address a single question (cancer detection, antibody quantification) or set of related questions (multiple mutation identification) in a narrowly defined specimen. For example, Paige Prostate was trained to detect prostate adenocarcinoma in needle prostate biopsies and is FDA approved for that diagnosis in that specimen type (19,20).

### What do we know about contaminants?

There is a robust literature around slide quality and artifacts in digital pathology (21–24). Recent work has shown that digitally mimicking artifacts including out-of-focus areas, threads, folds, marker, and crush artifact results in increasing rates of patch misclassification with more severe artifacts (21,24). Some countermeasures have been considered - inclusion of fields with imaging artifacts during training improves robustness to those artifacts at inference without compromising performance on pristine images (25). Pantanowitz et al. reported on a version of their Yottixel system to suggest the probable source tissue of floaters, although their system relies on pathologists to identify the tissue regions of concern (26).

### Systematic contaminants

Differences in procedure between institutions or over time may create non-random patterns of contamination. In placental pathology, guidelines support submission of membrane rolls and umbilical cord in the same block (27). However, our institution submits membrane and umbilical cord separately. Thus, a model trained using membrane roll slides from our institution would not be exposed to umbilical cord during training. If it were deployed at a site that co-submitted membranes and umbilical cord, the response to the cord is uncertain. Another example may be seen in prostate biopsy. Historically, most biopsies were performed trans-rectally, with a shift in the past decade to trans-perineal biopsies (28,29). Biopsy needles are designed to capture only prostatic tissue, but there is some risk of “pick-up” from the tissues transited. Thus, older prostate biopsies are more likely to contain fragments of large bowel, while newer biopsies are more likely to contain skin and subcutaneous tissue.

### Placenta, models

A diverse spectrum of placental abnormalities have been linked with a variety of fetal and neonatal outcomes (27,30). The breadth of these anomalies, high interobserver variability, and sparsity of perinatal pathology expertise motivate ongoing work in this field (31–34). Placenta is considered high risk as a destination for tissue contaminants due to its sponge-like architecture (14). Prior studies in the placenta have demonstrated classification and detection of normal and abnormal villous morphology (35–37), cell type and relationships (38), gestational age (39), and decidual arteriopathy (40). For this study, we chose to examine three models in placental pathology and physiology:

Decidual arteriopathy (DA) is a spectrum of abnormalities in decidual vessels associated with gestational hypertension and preeclampsia (27). DA is part of the maternal vascular malperfusion group of diagnoses. DA, specifically mural hypertrophy of membrane arterioles has been associated with SARS-CoV-2 infection in pregnancy, though concerns have been raised about interobserver variability in this diagnosis (41–43). Previous work by Clymer et al. has demonstrated the identifiability of DA in whole slide images of the placental membranes (40).

The placenta undergoes a reproducible series of changes over gestation, correlating with the clinically estimated gestational age (GA). Accelerated maturation is associated with growth restriction and gestational hypertension, while delayed maturation may be associated with diabetes in pregnancy (44–46). We previously published a model to predict GA(39).

Several macroscopic lesions can be identified within the placental disc, including villous infarction, perivillous fibrin deposition (PVFD), and intervillous thrombi (IVT). Villous infarction are areas of interrupted maternal circulation associated with hypertension in pregnancy, preeclampsia, growth restriction, stillbirth, and risk of cerebral palsy.(47–51) Perivillous fibrin deposition (PVFD) may be focal, where it is thought to represent a reparative response to turbulent flow, or massive (MPVFD) where it can be associated with fetal growth restriction and stillbirth.(52–56) Intervillous thrombi (IVT), are foci of clotted blood in the intervillous space.

They have no clear clinical significance, but merit recognition due to their frequency and risk of confusion with infarcts and PVFD.(41,57,58) We recently developed a model to distinguish these three lesions from one another and from placental disc sections without macroscopic lesions (59).

These problems correspond to three of the most common tasks in pathology – finding a small area of abnormality, estimating a quantity based on gestalt, and classifying a known abnormality.

### Prostate, model

Cancer detection is a common paradigm in ML and models to identify prostate cancer are among those FDA approved. We developed a model to detect prostatic adenocarcinoma in needle biopsy specimens.

## MATERIALS AND METHODS

### Dataset

#### Placentas

Inclusion criteria were patients that underwent placental examination and reporting at our institution between 2011 and 2023 and had slides scanned as part of an ongoing digitization study (IRB: STU00214052). Each model used additional criteria as noted below. Placentas were examined and diagnoses rendered according to the Amsterdam criteria or precursor guidelines (27). Slides were digitized using a Leica GT450 scanner with a 40x objective magnification (0.263 microns per pixel). The 10x magnification layer was used for all studies. Pathology reports were obtained from the institutional electronic data warehouse (EDW) and processed using natural language processing (42). Patient and diagnosis information was stored in REDcap.

#### DA model

Additional inclusion criteria were patients delivering after 26 weeks, 0 days (27). DA cases were those with a clinical diagnosis of mural hypertrophy of membrane arterioles. Controls were those without this diagnosis. One slide containing membrane roll from each placenta was selected. A total of 2336 cases were used, split 70:15:15 between training, validation, and test sets with stratification by case or control. The model was trained using the Adam optimizer and Focal loss. Early stopping was used when the validation loss plateaued.

#### GA model

Additional inclusion criteria were patients delivering a singleton at 24 weeks, 0 days gestation or later with a clinical diagnosis of appropriate villous maturation for stated gestational age (39,60). All placentas meeting these criteria were used. Up to three slides containing non-lesional villous tissue were used per patient. 846 placentas were used, randomly split 70:10:20 between the training, validation, and test sets. Splits were performed stratified by gestational age. The model was trained using the Adam optimizer and Huber loss. Early stopping was used when the validation loss plateaued.

#### Macroscopic lesion model

Additional inclusion criteria were diagnosis of infarction, PVFD, or IVT (for cases) or none of those for controls. Exclusion criteria included other macroscopic lesions, such as infarction hematoma, chorangioma, or SARS-CoV-2 placentitis. One slide was used per placenta, either containing lesion (for cases) or non-lesional villous tissue (for controls). 833 cases were split 70:15:15 into training, validation, and test sets. The model was trained using the Adam optimizer and categorical crossentropy loss. Early stopping was used when the validation loss plateaued.

#### Prostate adenocarcinoma model

Inclusion criteria were patients with prostatic needle biopsies examined at our institution, with slides scanned as part of an ongoing research effort. The dataset included 1-3 H&E slides each from 2602 blocks representing 647 patients. The Gleason grade assigned to each block was retrieved, with blocks classified as cancer if any cancer was present and otherwise negative.

Cases were split 80% training: 20% test, stratified by the presence of cancer, with all slides from a single patient assigned to the same group. Training and validation were similar to the placenta models except that features were extracted at 20X magnification with patch size of 224x224 pixels and no overlap. The feature extractor used was ConvNeXtXLarge, which produces a feature vector with 2048 values. Hinge loss was used.

#### Model – conceptual

All the models were implemented using Python version 3.7.7 and TensorFlow version 2.9 (61) An overview of the model is shown in **Figure 1**. First, the slide or slides comprising each case is split into a set of smaller patches. Patches undergo feature extraction – the first step of any image ML pipeline. Features are fed into an attention subnetwork, which assigns an attention to each patch, and produces a pooled feature vector weighted to the most highly attended patches. The pooled vector is run through a fully connected subnetwork to produce the result. The attention subnetwork and fully connected subnetwork are trained to minimize error. All models use a batch size of 1.

**Figure 1:**
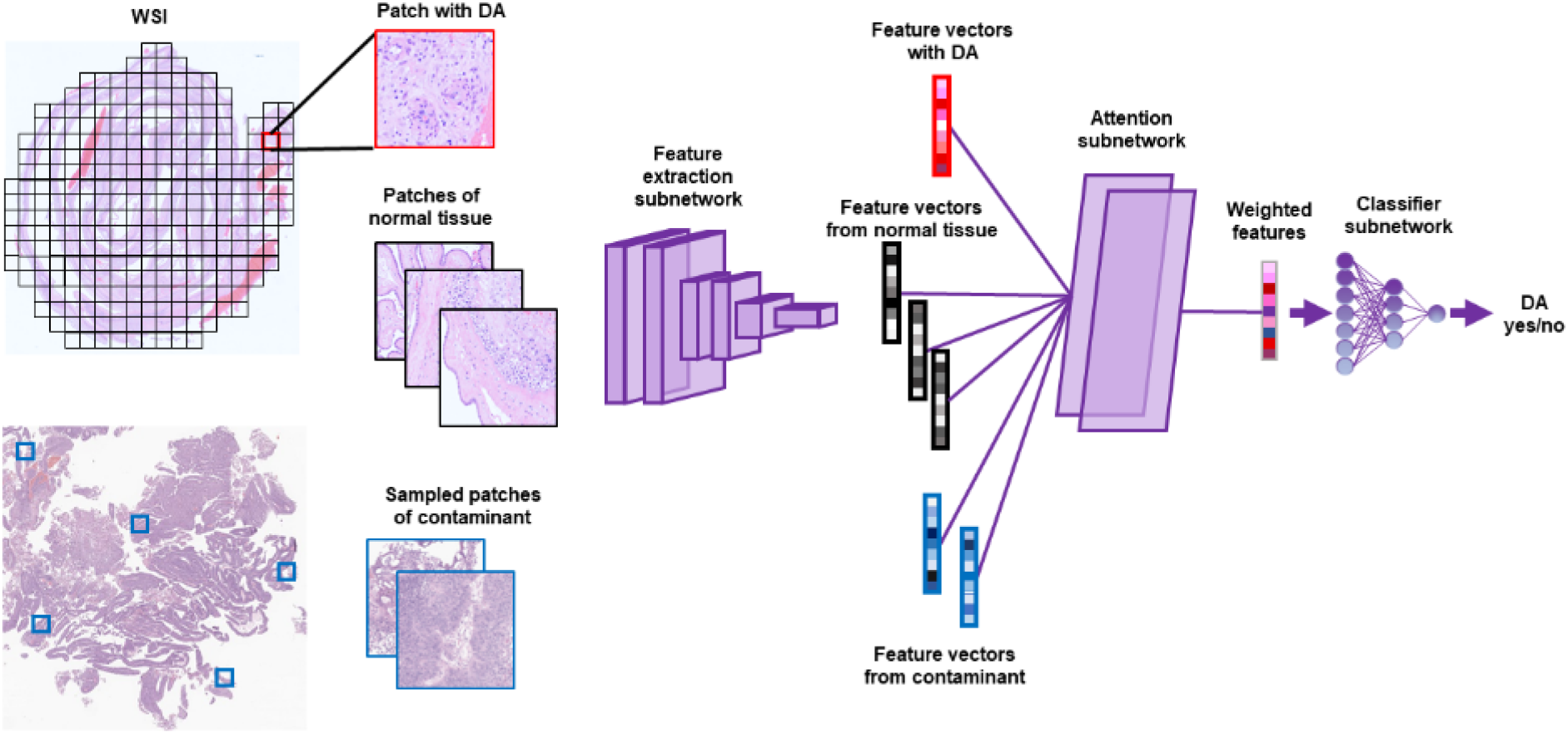
Overview of model and contaminants. Detection of DA is shown. Other diagnoses are conceptually similar. A whole slide image is split into *patches* (small squares), which may (red outline) or may not (black outline) have DA. The patches are submitted to a fixed feature extraction subnetwork, resulting in *feature vectors*. Feature vectors are submitted to the attention subnetwork which assigns an attention score to each feature vector. A *weighted feature* vector, representing a weighted average of the feature vectors is generated. The weighted features are submitted to a classifier subnetwork to produce a result. During training, the parameters of the attention and classifier subnetworks are varied to minimize error. To test the impact of tissue contaminants, random patches are sampled from the contaminant slide, feature extracted, assigned attention, and added to the weighted average.

#### Feature extraction

An image layer (10x for placenta, 20x for prostate) was split into patches (256x256 pixel for placenta, 224 × 224 pixel for prostate, **Figure 1**). Non-tissue patches were masked using Otsu’s method (62). Each patch is passed through a fixed feature extractor network, EfficientNetV2L(63), or ConvNeXtXLarge(64), each trained on ImageNet. After that, the feature vectors generated as ***NxD*** where ***N*** was the number of patches that were analyzed for each case and ***D*** was the number of features in each vector - 1280 for EfficientNetV2L, 2048 for ConvNeXtXLarge. Feature vectors were written as TensorFlow record files offline.

#### Attention subnetwork

The feature vectors obtained from the feature extraction were given to a trainable dense reducing dimensionality to a 512-dimensional feature. Then, to generate attention for each patch, we used the dot product of two parallel layers with 256 neurons, one with hyperbolic tangent activation and the other with sigmoid activation. Attentions for all examined patches are normalized to sum to 1 using a softmax function.

#### Classification subnetwork

The pooled feature vector is submitted to fully connected layers with appropriate activation. For determining class boundaries, individual patches are submitted to the classification network.

#### Contaminant slides

Contaminant slides were one each of low grade urothelial bladder tumor retrieved by transurethral resection (“bladder”), post-delivery non-adherent blood received with a placenta (“blood”), colonic mucosal biopsy showing a tubular adenoma (“colon”), fallopian tube fimbriae removed by salpingectomy for fertility control (“fallopian”), full thickness section of placental disc (“placenta”), hypertrophic prostate excised by holmium laser (“prostate”), excised skin with intradermal nevus (“skin”), small bowel resected after traumatic injury (“small bowel”), and umbilical cord cross sections (“umbilical”). Contaminant slides were reviewed to ensure they themselves were not contaminated, but not otherwise selected or resampled. Contaminant slides were scanned using the same Leica GT450 scanner as placentas and prostate biopsies and underwent the same feature extraction with the same magnification, tile size, and overlap. To add contaminant to a patient slide, a random subset of contaminant patches was selected and appended to the set of feature vectors from the placenta. The quantity of contaminant was varied, with 10% of contaminant indicating 10 patches of contaminant added for every 100 patches of relevant tissue. Unlike typical image corruption paradigms, the patient tissue patches are not altered or removed.

#### Interpretation model by using tSNE plots

T-distributed Stochastic Neighbor Embedding (tSNE) is used to view high-dimensional data in a lower-dimensional space (embedded space). In this study, we used tSNE from sklearn 1.2.0(65) for quantitative model performance analysis to reveal how attention and classification of individual patches clusters. tSNE parameters perplexity and random were 30 and 0 respectively for two dimensions of the embedded space.

## RESULTS

### Multiple contaminants interfere with detection of decidual arteriopathy (DA)

We trained a model to detect DA. At baseline, the model showed sensitivity of 0.6, specificity of 0.88, AUC of 0.81, and balanced accuracy of 0.74 (**Figure 2, Supplementary Table 1**). Slides of normal membranes far outnumber normal DA cases in this dataset, so balanced accuracy is a more representative measure. Each contaminant decreased balanced accuracy somewhat with prostate showing the largest impact. Adding 1% prostate tissue reduced the balanced accuracy from 0.74 to 0.69 +/- 0.01. 10% prostate tissue reduced accuracy to almost chance levels. Surprisingly, different contaminants resulted in different types of error. Small bowel, fallopian tube, and bladder resulted in false positive calls, while blood, umbilical cord, and prostate resulted in false negatives.

**Figure 2.**
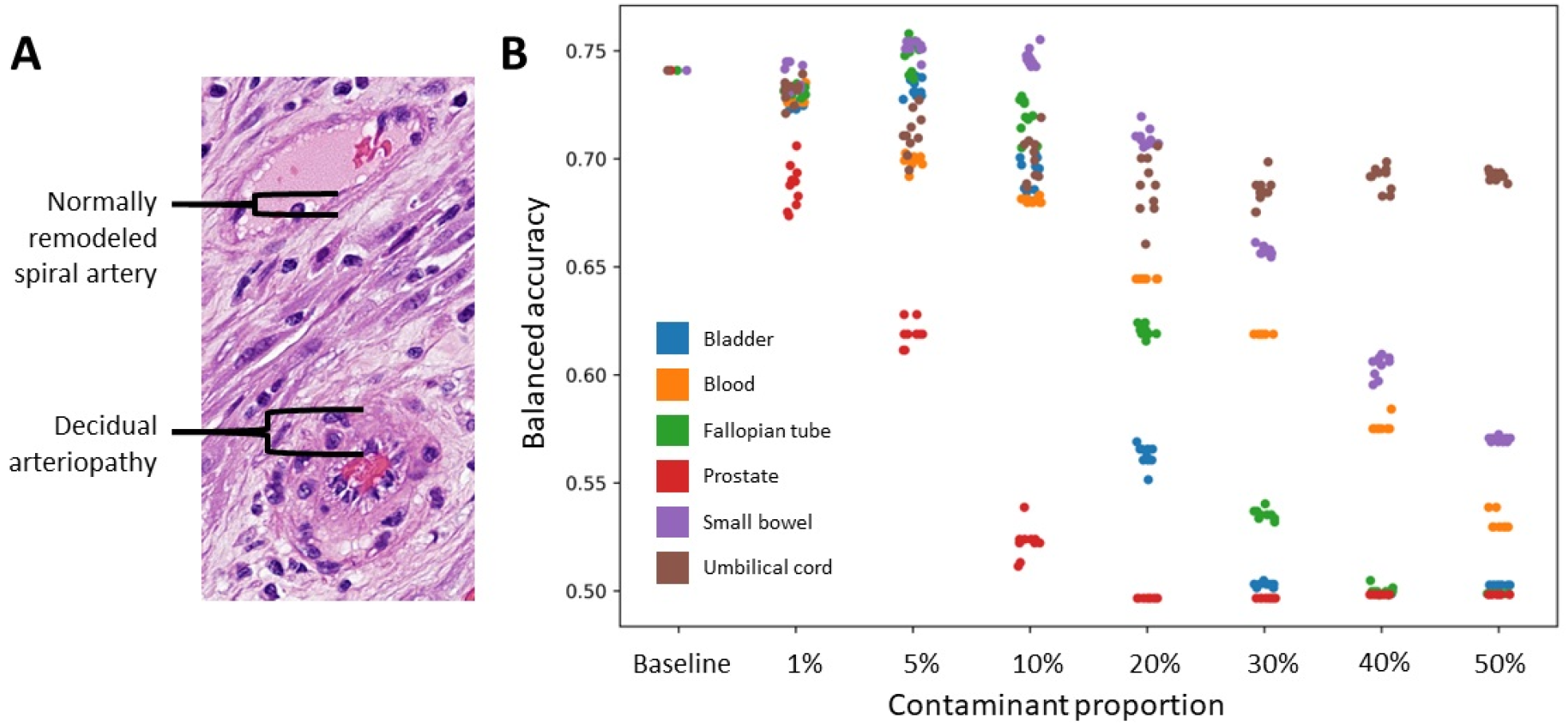
Multiple contaminants impair balanced accuracy in detecting DA. A normally remodeled spiral artery (top) and spiral artery with DA, specifically mural hypertrophy of membrane arterioles (bottom) are shown for illustration (A). Addition of even 1% of prostate tissue causes noticeable decrements in balanced accuracy, while bladder, fallopian tube, and blood at higher proportions also bring the accuracy near to or at chance level (0.5, B).

### Contaminants decrease accuracy of gestational age (GA) estimation

We developed a model to estimate gestational age by examination of placental villi. At baseline, the model shows strong performance, with an R^2 of 0.75 and a mean absolute error (MAE) – the average difference between the estimated gestational age and the chronologic gestational age - of 1.66 weeks (**Figure 3, Supplementary Table 2**). Adding contaminants resulted in increased MAE, with bladder causing errors of 2.37 weeks at 5% contaminant and 3.36 weeks at 10% contaminant. Interestingly, the impact on estimated GA was monotonous, such that even though the MAE with 30% bladder was 6.87 weeks, the R^2^, which depends primarily on the order of data points, only fell to 0.72.

**Figure 3.**
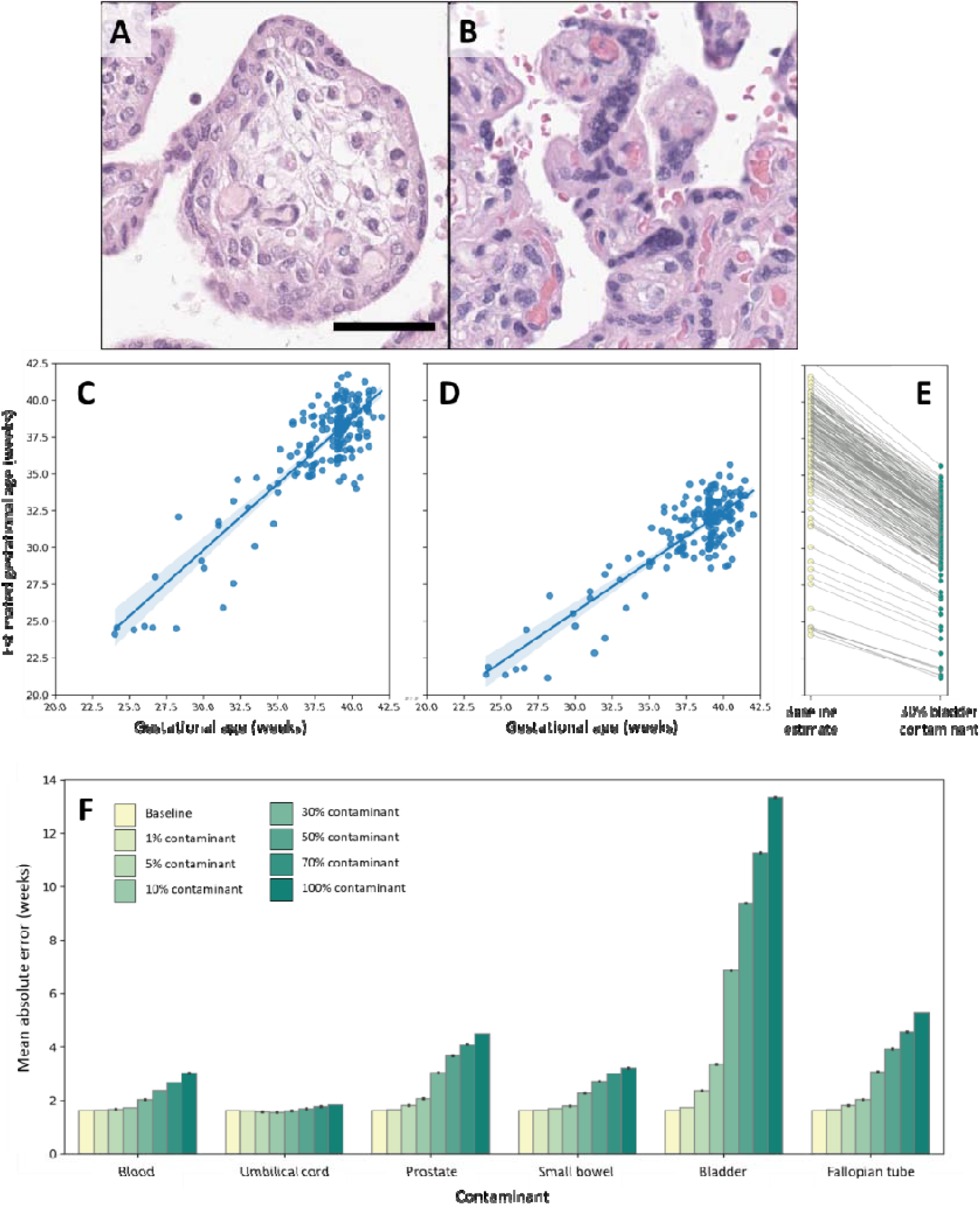
Impact of contaminants on estimation of GA. Placental villi undergo a reproducible series of morphologic changes between 24 (**A**) and 42 (**B**) weeks gestation. Villi are narrower with denser stroma, more exteriorized capillaries, and increased syncytial knots. Our baseline model is highly accurate with a mean absolute error (MAE) or 1.66 weeks (**C**). Addition of contaminant, such as 30% bladder (example shown in **D**) results in sharply higher MAE. The decrease appears monotonous, with each case showing a similar decrease in estimated GA (**E**), which results in a misleadingly preserved correlation coefficient (R^2^). While bladder clearly shows the highest impact, other tissues including blood, prostate, small bowel, and fallopian tube also increase error, albeit at higher proportions (**F**). Umbilical cord did not result in significantly increased errors. Scale bar 50μm.

**Figure 4:**
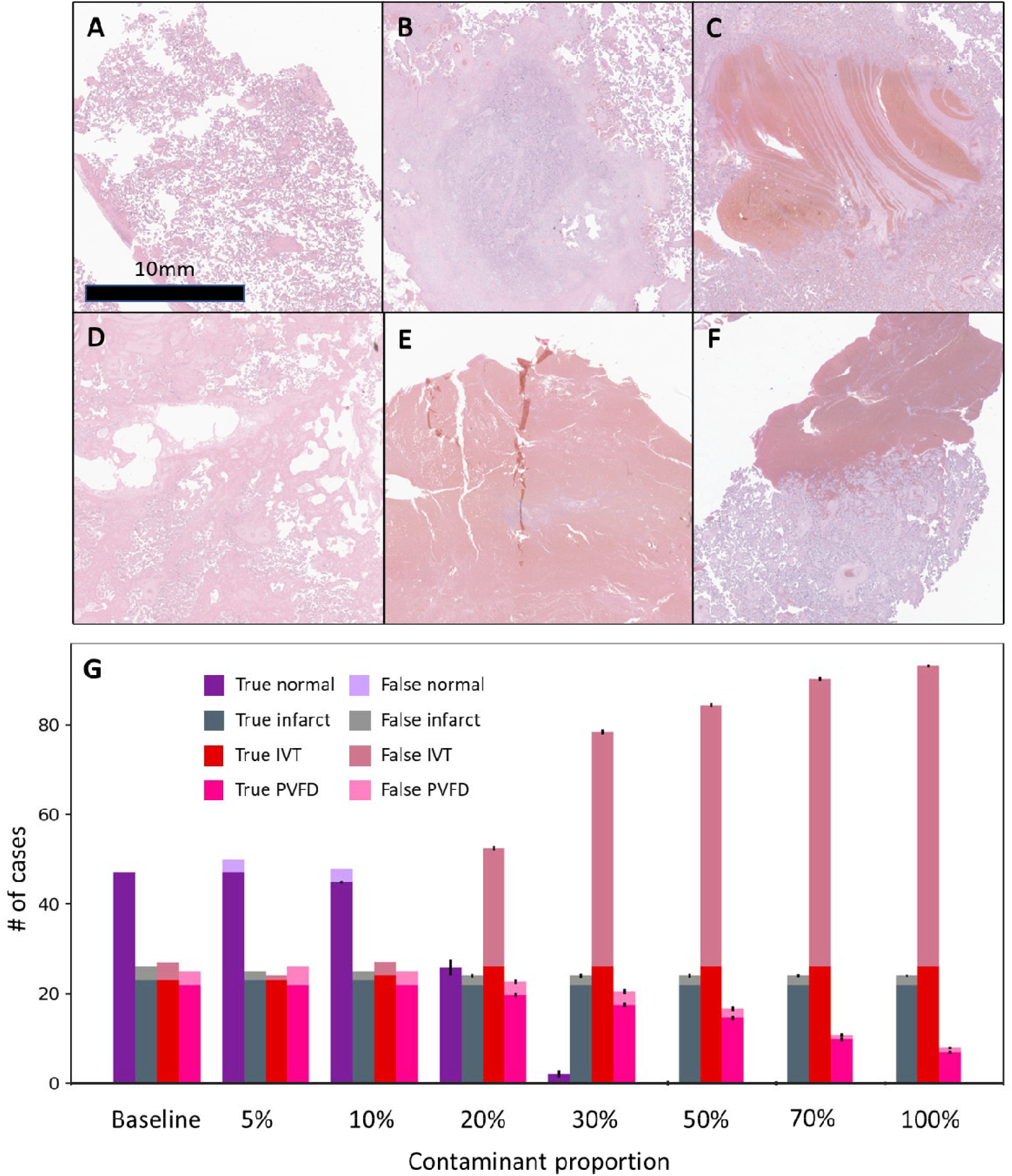
Blood causes misclassification of macroscopic placental lesions. Paradigmatic examples of normal placenta (A), infarct (B), IVT (C), and PVFD (D). A section of fresh blood was used as contaminant (E). Note Lines of Zahn as a critical distinguishing morphology (B vs. E). In clinical practice, areas of unclotted blood may be submitted incidentally with normal placenta sections, causing misclassification as IVT (F). The lesion classification model is highly accurate at baseline, with balanced multiclass accuracy of 0.89. With increasing amounts of blood contaminant, normal and PVFD cases are progressively misclassified as IVT. At baseline, 4 cases are misclassified as IVT. At 20% blood, 26.5+/-2.0 cases are misclassified as IVT. Scale bar: 10 mm.

### Blood contaminant causes misclassification of macroscopic placental tissue as intervillous thrombi

We tested the impact of tissue contaminants on a model that classifies placental slides as either villous infarction (infarct), intervillous thrombus (IVT), perivillous fibrin deposition (PVFD) or none of the above (normal). At baseline, the model has an accuracy of 0.89. Bladder, fallopian tube, prostate, small bowel, and umbilical cord had minor impacts (**Supplemental Table 3**).

However, addition of blood caused misclassification of normal and, to a lesser extent, PVFD, slides as IVT. 4 cases were misclassified as IVT at baseline, reaching 26.5+/-2.0 cases at 20% contaminant and 64.2 +/- 0.4 at 70% (n=10 replicates). Infarcts, actual IVT and, to a lesser extent, PVFD remained accurately classified, so balanced accuracy poorly reflects the error, falling only from 0.894 at baseline to 0.891 at 20% contaminant, with a larger drop to 0.60 at 70% contaminant.

### Contaminants cause false positives in a prostate cancer detection model

We developed a prostate cancer detection model with a baseline accuracy of 0.923 and AUC of 0.954 (**Figure 5, Supplementary Table 4**). The model was extremely sensitive to bladder contaminants, which resulted in high rates of false positives. Other contaminants, including colon and fallopian tube resulted in lower magnitude errors.

**Figure 5.**
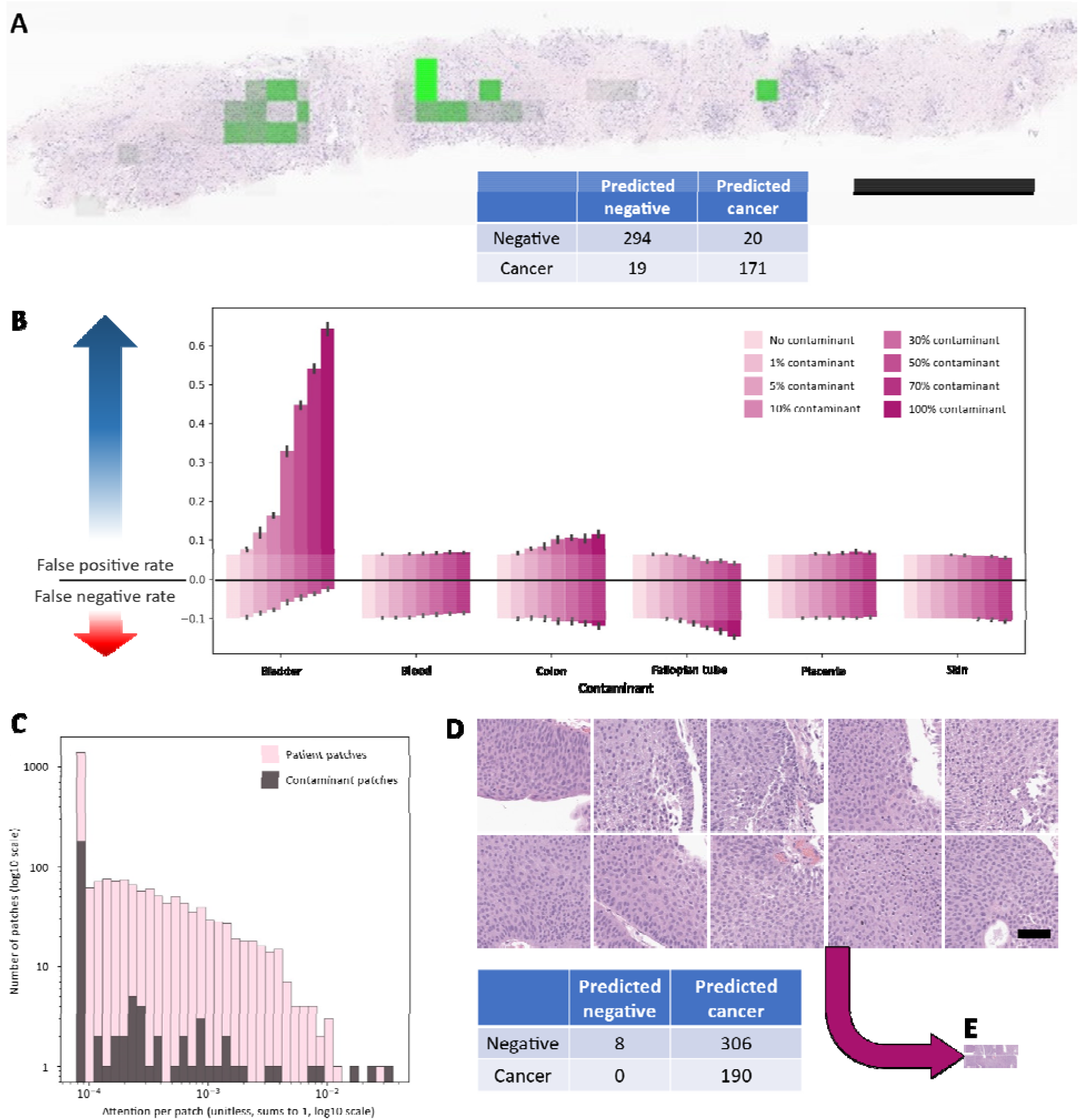
Prostate cancer detection model. We developed a model to detect prostate carcinoma in needle biopsies, which attends to areas of cancer and results in high block-level accuracy (**A**, scale bar 1 mm). Adding bladder, resulted in marked increases in the false positive rate, from 20/314 at baseline to 51+/-2.7 at 10% bladder contaminant and 197+/-7.9 when the amount of added bladder equaled the amount of prostate tissue (100% contaminant, **B**, n=10 replicates). Patch level attention of one selected case with 10% bladder contaminant (**C**) showed the expected power law distribution. Surprisingly, 3 of the 4 most highly attended patches were contaminant, rather than patient tissue. We identified the 10 most highly attended patches of the contaminant bladder slide (**D**, scale bar 20μm) all of which show unremarkable low grade bladder tumor. Adding these 10 patches to each prostate case results in 306/316 false positives. These patches represent a minute quantity of tissue (**E**, patches from **D** resized to same scale as **A**).

#### All contaminants distract attention in all models

We measured the proportion of attention given to each contaminant by each model (**Figure 6**). Unsurprisingly, increasing amounts of contaminant received increasing amounts of attention. More surprisingly, the level of attention given to contaminant tissues often exceeded, patch for patch, that given to patient tissue. This represents a significant failure of the attention mechanism, even if errors did not result. High attention did not always correlate with high rates of error. For example, umbilical cord was among the most highly attended tissues by the GA model, particularly at lower proportions, but it did not result in errors.

**Figure 6:**
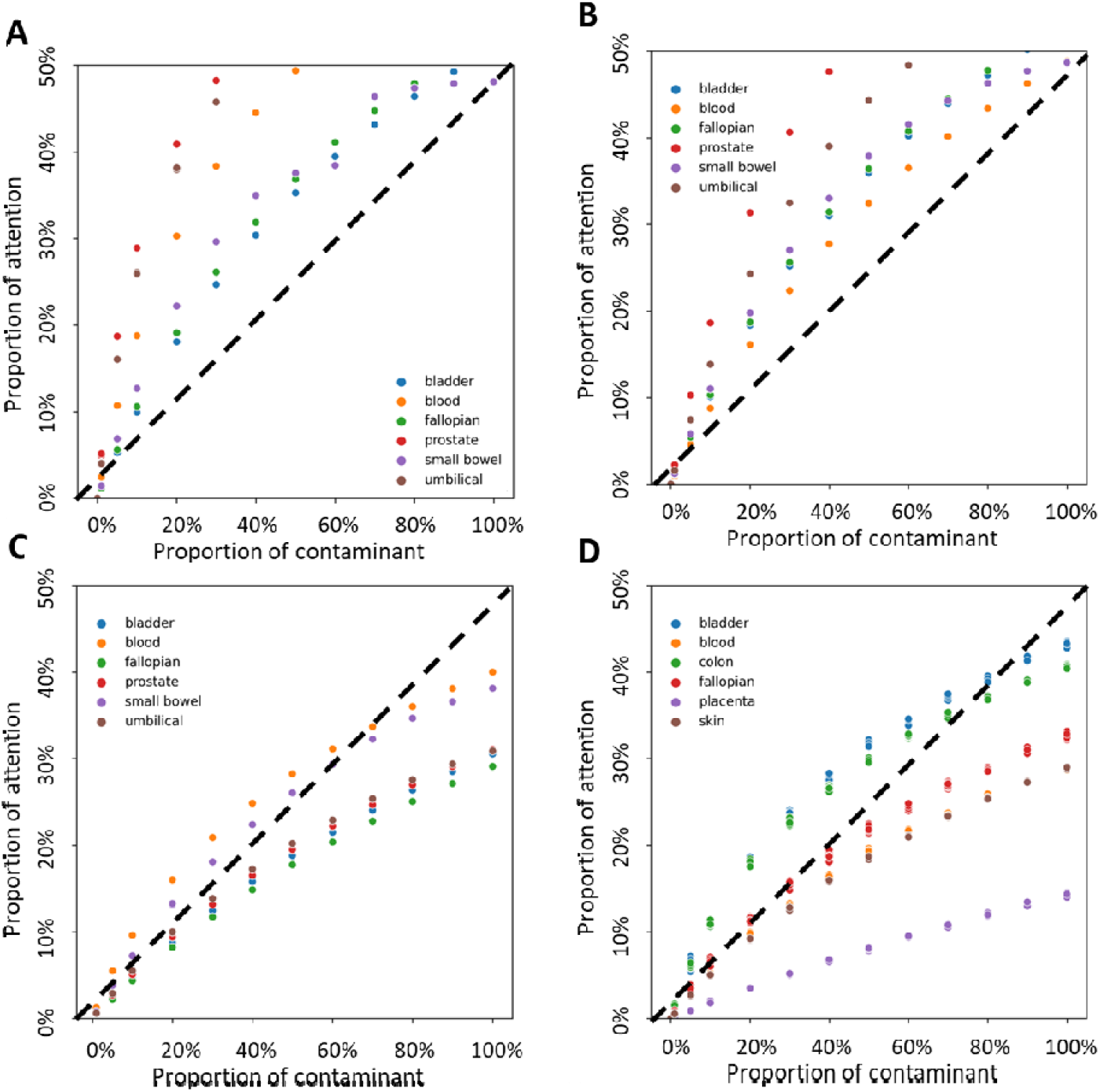
All contaminants distract attention. We measured the proportion of attention given to contaminant patches as a function of the proportion of contaminant added for models of DA (**A**), GA (**B**), macroscopic lesions (**C**), and prostate adenocarcinoma (**D**). The dashed line represents attention *at par*, where the average patch of contaminant receives as much attention as the average patch of patient tissue. For example, 20% contaminant represents 20 patches of contaminant for each 100 patches of patient tissue. Therefore, the contaminant represents _______ ^__^ of the patches present. Every contaminant received some attention. Multiple contaminants in DA, GA, lesions, and prostate adenocarcinoma being attended above par.

### Contaminants occupy a distinct location in embedded feature space feature space and divert pooled features from the decision boundary

To track the impact of contaminants on model decisions, we, visualized the feature space using tSNE (**Figure 7**). The patches classified as positive or negative occupy distinct regions. By applying the same transformation to the pooled feature vector, we can see where the model places the specimen overall. In a case that shifts from true negative to false positive at a low contaminant proportion, the feature vector is close to the apparent boundary between the positive and negative instances. Adding a relatively small amount of contaminant shifts the pooled feature vector in feature space, consistent with the changed slide-level interpretation.

**Figure 7.**
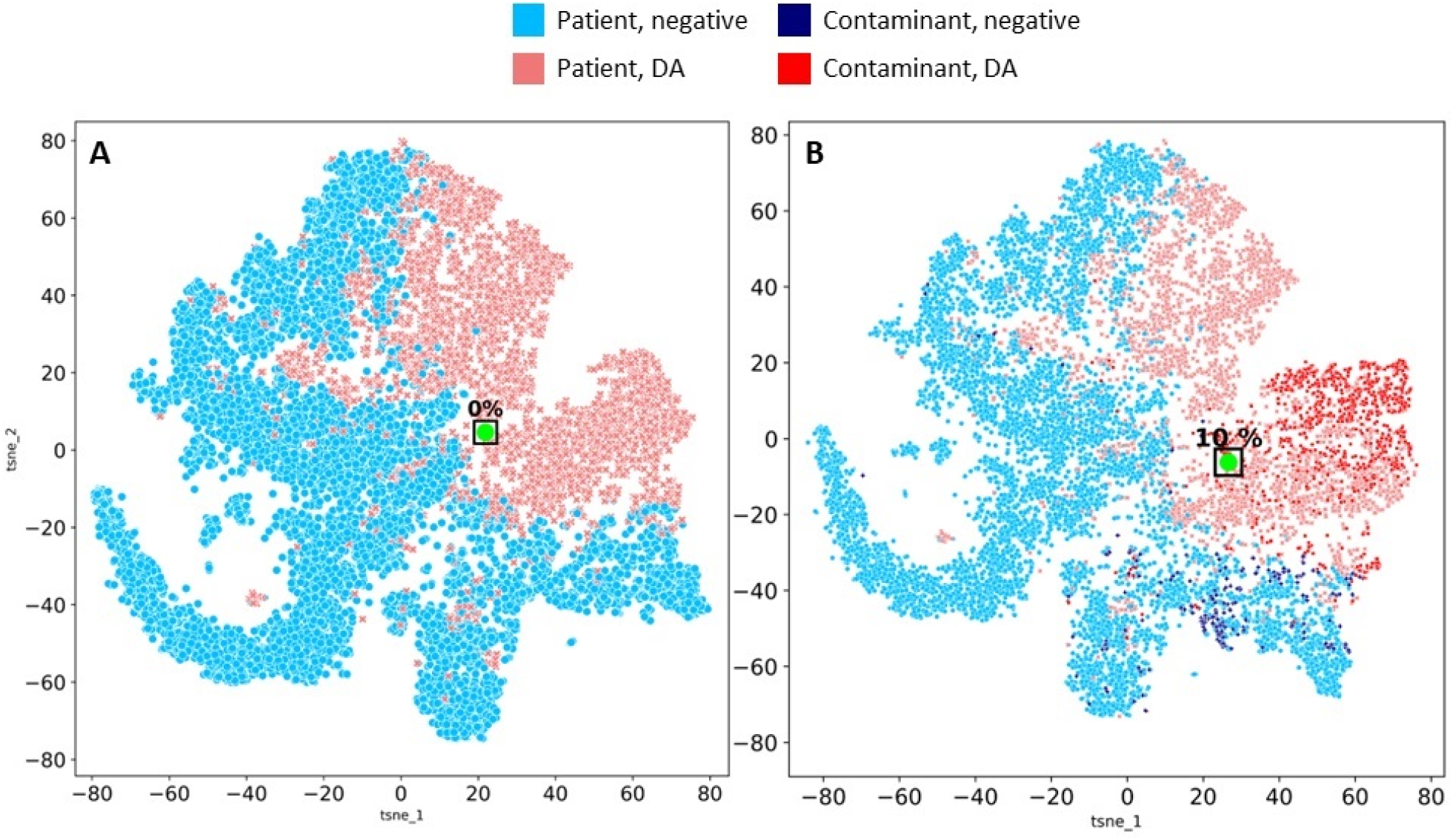
impact of contaminants in features space. Feature maps for one slide, a true negative at baseline (A) that becomes a false positive when 10% bladder contaminant is added (B). Each point represents one patch, transformed into a feature vector and then plotted in 2-dimensional space using tSNE. Patches are classified as DA (rose) or not DA (light blue), which form largely distinguishable. The pooled feature vector, representing a weighted average, is marked in green. Added contaminants are largely classified as classified as DA (bright red), with a few not DA (dark blue). Adding contaminant causes the pooled feature vector to more clearly cluster with the positive instances, resulting in the false positive call.

## DISCUSSION

### Recitation

We report four whole-slide ML models developed with contemporary techniques. The models encompass tissues including placental membrane, disc, and prostate core and tasks including detection, classification, and regression. Each model shows degraded performance when tissue contaminants are added to previously well-classified data. Each model attends to contaminating tissue, often at-or-above the level shown to patient tissue. Contaminating tissue induces errors by altering the attention distribution within feature space and redirecting the pooled feature vector across the decision boundary.

### Comparison

In the broadest sense, all machine learning models perform reliably when given data like that seen in training and perform unpredictably when given data from outside that distribution. More narrowly, this work falls within the digital pathology ML and computer vision topics on image corruptions or visual artifact pathology (21–24). Our findings contribute to this literature with a few key differentiations. Our focus on the attention mechanism identifies a key weakness for whole-slide models. Using added contaminant, rather than distorting or obscuring the underlying image, guarantees that the original diagnostic information is preserved intact. Tissue contaminants themselves are a recognized problem in the pathology quality literature but have not attracted broader attention (11,14).

False positives due to tissue contaminants have been previously reported in ML cancer detection tasks. Liu et al. reported one floater (of 80 slides) causing false positive metastatic breast cancer diagnoses in the CAMELYON dataset, although they did not further consider the implications (66). Similarly, Campanella show a false-positive lymph node due to a benign papillary inclusion in a lymph node – an example of a biological systemic contaminant (67).

Naito et al. reported only 1 false positive case of pancreatic cancer in their test set (of 34 slides), however that sole false positive was due to a “pick-up” of gastric tissue transited during the endoscopic biopsy (68). This form of systemic contamination is described as “ever present” in pathology textbooks, so the true risk may be higher (69). These previous publications suggest that false positive cancer detection due to tissue contaminants may be endemic.

Our methods – altering patient data to test robustness – are similar to those seen in adversarial attacks (70). In the digital pathology literature, these studies have taken the form of alterations to individual patches that are imperceptible to human observers, yet induce incorrect responses from models (10,71,72). Our work is similar, in that the alterations – subtle pixel-level alterations or tissue contaminants – are ignored by trained human observers. However, there are key differences. We focus on attention-level mechanisms, leaving the original data completely unchanged. Our ‘attack’ does not rely on access to the model or model architecture. Most significantly, while adversarial attack invokes the (so far unobserved) possibility of bad actors gaining access to digital pathology images and making targeted alterations, tissue contaminants are already inside the building.

### Attention is a core model mechanism and key model output

Saliency mapping, such as GradCAM, has been used to demonstrate shortcuts taken by models, such as using hospital-specific markings to classify chest X-rays as COVID-19 pneumonia, rather than examination of the lungs (73). The case in digital pathology is similar, but the problem is more acute. First, while GradCAM and other saliency mechanisms are post-hoc explanations, attention in whole-slide models is central to how the model produces output. Second, in a computer assisted diagnosis paradigm, pathologists will primarily be working with visualizations of the attention maps, rather than the final prediction. In that sense, the attention is a much more important output than the prediction. If the model is generating accurate slide-level predictions but attending to irrelevant tissue, pathologists will question the credibility of the model.

Those doubts are well-founded. By attending to contaminant tissue, the models demonstrate that they do not encode the underlying biological phenomena, in this case that pathologies are tissue specific. Prostate cancer does not arise in the bladder. Decidual arteriopathy arises only in the decidua – not the prostate. While it is shocking that the GA model attends to the average patch of prostate twice as much as it attends to the average patch of placenta, any significant attention given to contaminants demonstrates a critical failure.

### Considering contaminants – source and quantity

Our contaminants are a convenience sample of tissues commonly encountered in surgical pathology practice and include some of those – colon, bladder, prostate, and placenta – seen as highest risk for producing or capturing contaminants (12,14). We tested only one slide per tissue source, so different diagnoses (e.g. high-grade vs. low grade bladder cancer) or different slides with the same diagnosis are likely to give different results. While we acknowledge this limitation, we would argue that the arising uncertainty proves our point – model performance is worryingly unpredictable in this circumstance.

Insofar as our results are representative, bladder and prostate seem to be high risk tissues in general, but this is likely dependent on the ML task. Unclotted blood is commonly received with placental specimens and submitted for histology (see **Figure 4F**). The distinction between this blood and true thrombi, indicated by the Lines of Zahn, is a core pathologist skill (74). The volume of blood thus submitted is often large, easily representing 1/3 of tissue present (i.e. 50% contaminant), sufficient to induce errors in multiple placental classifiers. Umbilical cord sections are often submitted with membrane rolls, usually at a ratio of 1 cord section: 1 membrane roll – 100% contaminant. At that level, the cord sections cause a small but meaningful decrease in DA accuracy (**Figure 2**). Fewer institutions submit cord sections with placental disc, but if they do, it seems unlikely to alter their GA estimation (**Figure 3**). The switch from transrectal to transperineal prostate biopsy, in addition to its other benefits, would also seem likely to reduce the risk of false negative and false positive results (**Figure 5**). The size of sporadic contaminants is likely to be smaller, reported 1 mm^2^ on average (13). This represents a very small proportion of a placental disc section, but a larger proportion of a prostate needle biopsy. While the effect size increases with the proportion of contaminants, errors were detectable at 1% contaminant for DA, GA, and prostate.

### Next Steps - what can be done

The data presented in this paper are sufficient to establish tissue contaminants as a risk to whole-slide learning models in modern digital pathology. We do not aim for a complete accounting or explanation of tissue contaminants, nor are there simple answers to address this problem. Use of humans or different ML models to ‘pre-screen’ slides for contaminants suggests itself as a solution but is fraught. If ML models are needed to increase efficiency in the face of increasing specimens and a declining pathologist workforce, adding human intervention is self-defeating (75).

My concern with adding humans is that we need (really truly need) AI to allow a diminishing number of pathologists to keep up with an expanding number of specimens. My concern with building a model for contaminants is that it needs to be able to recognize all tissues with all different pathologies. It also needs to be able to recognize uncommon variants of pathology at your site of interest and not exclude them. If your screening model is that good, you don’t need the single-purpose model. Regarding the use of a pre-screening ML model to identify contaminants - the diversity of tissues and their pathologies in the human body necessitates a very advanced model to distinguish contaminants from atypical presentations of the disease of interest. Such an advanced prescreener would obviate the need for specialized single-organ models.

There are some findings that suggest future actions for pathologists, decision makers, and ML practitioners. 1) An ML transformation of pathology may require a renewed emphasis within the lab on reducing tissue contaminants. 2) clinical decisions should continue be made with a human-in-the-loop. As described in the introduction, human pathologists recognize tissue contaminants as a basic skill – that expertise should be used. 3) Model vulnerability to common tissue contaminants should be assessed. 4) Work is needed to identify countermeasures to tissue contaminant-induced error. Statistical approaches could be used for all out-of-distribution data, while training against high-risk contaminants may blunt their impact.

## Data Availability

Whole slide image data are available after execution of a data use agreement with Northwestern University.

## Funding / Acknowledgements / Conflict of interest

JAG is supported by NIBIB K08EB030120 and the Walder Foundation Fund to Retain Clinician Scientists. LADC is supported by R01LM013523, and U01CA220401. REDCap and other key infrastructure supported by UL1TR001422.

The authors state that no conflict of interest exists.

